## Supplementary Material for "Seroprevalence of anti-SARS-CoV-2 IgG antibodies in Tyrol, Austria: Updated analysis involving 22,607 blood donors covering the period October 2021 to April 2022"

Seekircher and Siller et al.

### Supplementary tables

#### Table S1. STROBE checklist.

| | Item No | Recommendation | Page No |
|---|---|---|---|
| Title and abstract | 1 | (*a*) Indicate the study's design with a commonly used term in the title or the abstract | 1 |
| | | (*b*) Provide in the abstract an informative and balanced summary of what was done and what was found | 1 |
| **Introduction** | | | |
| Background/ rationale | 2 | Explain the scientific background and rationale for the investigation being reported | 1-2 |
| Objectives | 3 | State specific objectives, including any prespecified hypotheses | 2 |
| **Methods** | | | |
| Study design | 4 | Present key elements of study design early in the paper | 2-3 |
| Setting | 5 | Describe the setting, locations, and relevant dates, including periods of recruitment, exposure, follow-up, and data collection | 2 |
| Participants | 6 | (*a*) Give the eligibility criteria, and the sources and methods of selection of participants. Describe methods of follow-up | 2 |
| | | (*b*) For matched studies, give matching criteria and number of exposed and unexposed | NA |
| Variables | 7 | Clearly define all outcomes, exposures, predictors, potential confounders, and effect modifiers. Give diagnostic criteria, if applicable | 2-3 |
| Data sources/ measurement | 8* | For each variable of interest, give sources of data and details of methods of assessment (measurement). Describe comparability of assessment methods if there is more than one group | 2 |
| Bias | 9 | Describe any efforts to address potential sources of bias | 2-3 |
| Study size | 10 | Explain how the study size was arrived at | 2 |
| Quantitative variables | 11 | Explain how quantitative variables were handled in the analyses. If applicable, describe which groupings were chosen and why | 2-3 |
| Statistical methods | 12 | (*a*) Describe all statistical methods, including those used to control for confounding | 2-3 |
| | | (*b*) Describe any methods used to examine subgroups and interactions | 2-3 |
| | | (*c*) Explain how missing data were addressed | 2-3 |
| | | (*d*) If applicable, explain how loss to follow-up was addressed | NA |
| | | (*e*) Describe any sensitivity analyses | 2-3 |
| **Results** | | | |
| Participants | 13* | (a) Report numbers of individuals at each stage of study—eg numbers potentially eligible, examined for eligibility, confirmed eligible, included in the study, completing follow-up, and analyzed | 3-6 |
| | | (b) Give reasons for non-participation at each stage | 3-6 |
| | | (c) Consider use of a flow diagram | NA |
| Descriptive data | 14* | (a) Give characteristics of study participants (eg demographic, clinical, social) and information on exposures and potential confounders | 3, Table 1 |
| | | (b) Indicate number of participants with missing data for each variable of interest | Table 1 |
| | | (c) Summarize follow-up time (eg, average and total amount) | Table 1 |
| Outcome data | 15* | Report numbers of outcome events or summary measures over time | 3-6 |
| Main results | 16 | (a) Give unadjusted estimates and, if applicable, confounder-adjusted estimates and their precision (eg, 95% confidence interval). Make clear which confounders were adjusted for and why they were included | 3-6 |
| | | (b) Report category boundaries when continuous variables were categorized | 4-6 |
| | | (c) If relevant, consider translating estimates of relative risk into absolute risk for a meaningful time period | NA |
| Other analyses | 17 | Report other analyses done—eg analyses of subgroups and interactions, and sensitivity analyses | 3-6 |
| **Discussion** | | | |

| | | | |
|---|---|---|---|
| Key results | 18 | Summarize key results with reference to study objectives | 7 |
| Limitations | 19 | Discuss limitations of the study, taking into account sources of potential bias or imprecision. Discuss both direction and magnitude of any potential bias | 8 |
| Interpretation | 20 | Give a cautious overall interpretation of results considering objectives, limitations, multiplicity of analyses, results from similar studies, and other relevant evidence | 7-8 |
| Generalizability | 21 | Discuss the generalizability (external validity) of the study results | 7-8 |
| **Other information** | | | |
| Funding | 22 | Give the source of funding and the role of the funders for the present study and, if applicable, for the original study on which the present article is based | 9 |

**Table S2. Seroprevalences in all participants (principal analysis), in participants with repeated donations since July 2017, and in all participants with age and sex standardization across the total population of Tyrol, Austria.**

| Month | % Seropositive (95% confidence interval) | | |
|---|---|---|---|
| | All participants (n = 22,607) | Participants who had already donated blood leading up to the study* (n = 17,764) | All participants (n = 22,607) with age and sex standardization across the total population of Tyrol† |
| October 2021 | 84.9 (83.8–86.0) | 87.0 (85.8–88.1) | 85.3 (84.2–86.4) |
| November 2021 | 88.7 (87.6–89.6) | 90.2 (89.1–91.2) | 88.6 (87.6–89.7) |
| December 2021 | 91.3 (90.3–92.3) | 91.3 (90.2–92.3) | 91.3 (90.3–92.3) |
| January 2022 | 93.5 (92.8–94.2) | 94.1 (93.3–94.9) | 93.4 (92.6–94.2) |
| February 2022 | 95.3 (94.5–96.0) | 95.7 (94.9–96.5) | 95.2 (94.3–96.0) |
| March 2022 | 95.9 (95.2–96.5) | 96.2 (95.5–96.9) | 95.9 (95.2–96.5) |
| April 2022 | 95.8 (94.9–96.4) | 95.7 (94.8–96.5) | 95.9 (95.1–96.6) |

*To define this subgroup of the study population, the period from July 2017 to study baseline was considered. †Direct age and sex standardization was applied by using age (categories 18-30, >30-40, >40-50, >50-60, >60-70 years) and sex structured data of the population of the Federal State of Tyrol in Austria as standard population as of 1 January 2022 from the Statistik Austria.

#### Supplementary figures

**eFigure 1. Shift of vaccinated and unvaccinated participants across categories of anti-S IgG antibody levels in Binding Antibody Units per milliliter between first and latest available follow-up measurement between October 2021 and April 2022.**

**A Vaccinated at first and latest available follow-up measurement**

| First measurement \ Follow-up measurement | Seronegative | <500 | 500–<1000 | 1000–<2000 | 2000–<3000 | ≥3000 | Row total |
|---|---|---|---|---|---|---|---|
| Seronegative | 2 (11.1%) | 4 (22.2%) | 2 (11.1%) | 5 (27.8%) | 3 (16.7%) | 2 (11.1%) | 18 |
| <500 | 0 (0.0%) | 157 (14.2%) | 229 (20.7%) | 245 (22.2%) | 121 (11.0%) | 352 (31.9%) | 1104 |
| 500–<1000 | 0 (0.0%) | 48 (13.6%) | 64 (18.1%) | 76 (21.5%) | 42 (11.9%) | 123 (34.8%) | 353 |
| 1000–<2000 | 0 (0.0%) | 31 (9.9%) | 57 (18.2%) | 66 (21.0%) | 48 (15.3%) | 112 (35.7%) | 314 |
| 2000–<3000 | 0 (0.0%) | 7 (4.7%) | 34 (22.7%) | 52 (34.7%) | 19 (12.7%) | 38 (25.3%) | 150 |
| ≥3000 | 0 (0.0%) | 5 (1.8%) | 26 (9.6%) | 59 (21.7%) | 54 (19.9%) | 128 (47.1%) | 272 |

**B Unvaccinated at first and latest available follow-up measurement**

| First measurement \ Follow-up measurement | Seronegative | <500 | 500–<1000 | 1000–<2000 | 2000–<3000 | ≥3000 | Row total |
|---|---|---|---|---|---|---|---|
| Seronegative | 101 (51.8%) | 88 (45.1%) | 6 (3.1%) | 0 (0.0%) | 0 (0.0%) | 0 (0.0%) | 195 |
| <500 | 1 (0.8%) | 107 (84.3%) | 10 (7.9%) | 5 (3.9%) | 1 (0.8%) | 3 (2.4%) | 127 |
| 500–<1000 | 0 (0.0%) | 2 (100.0%) | 0 (0.0%) | 0 (0.0%) | 0 (0.0%) | 0 (0.0%) | 2 |
| 1000–<2000 | 0 (0.0%) | 0 (0.0%) | 0 (0.0%) | 0 (0.0%) | 0 (0.0%) | 0 (0.0%) | 0 |
| 2000–<3000 | 0 (0.0%) | 0 (0.0%) | 0 (0.0%) | 0 (0.0%) | 0 (0.0%) | 0 (0.0%) | 0 |
| ≥3000 | 0 (0.0%) | 0 (0.0%) | 0 (0.0%) | 0 (0.0%) | 0 (0.0%) | 1 (100.0%) | 1 |

Seronegativity corresponds to anti-S IgG levels <7.1 Binding Antibody Units per milliliter. The intensity of the cell color reflects the row percentage of the cell. (A) The analysis involved 2211 participants who were vaccinated with at least one dose of any vaccine against SARS-CoV-2 at both measurements. (B) The analysis involved 325 participants who were unvaccinated at both measurements.
